# A transformer model explaining mechanisms of drug therapeutic and adverse effects

**DOI:** 10.64898/2026.05.11.26352917

**Authors:** Jianfeng Ke, Rachel Melamed

## Abstract

Understanding which disease genes are altered by a drug can provide insight into the biology of effect, help us understand adverse drug effects, and suggest new drug uses. Here, we build on our model Draphnet in a new formulation with a similar goal. Draphnet was designed to explain drug therapeutic and side effects by learning a network connecting drugs to the disease genes they alter. Our new model, DraPhormer, has a similar goal but instead of relying on a linear model, learning of drug to gene connections uses a transformer model. DraPhormer integrates drug molecular data, disease genetics, and known drug effects on diseases, along with language models representing all of these entities. We show in simulations that DraPhormer can explain the genetic mechanisms of drug effects. Then, we present our design for incorporating drug and disease biology into the model. Finally, we benchmark the model’s ability to learn drug indications and side effects in real data.

## Introduction

A new paradigm in identifying drug therapeutic effects uses disease genetics to identify disease causal genes, and then prioritizes drugs impacting these genes(Nelson et al. 2015; King et al. 2019; Minikel et al. 2024). Similarly, drug adverse effects have also successfully been predicted using genes identified from genome-wide association studies (GWAS)(Nguyen et al. 2019; Minikel and Nelson 2023). But, such direct genetic evidence is sparse, and many drug effects have not been explained this way. We recently proposed an integrative model that we call Draphnet, that learns to predict a drug’s therapeutic and adverse effects via predicting the set of disease genes the drug impacts (Habib et al. 2024). The design is based on the premise that drugs that affect similar diseases do so by perturbing similar sets of genes linked to these diseases. This has the potential to take advantage of weaker signals, including the genetic similarity between pairs of diseases(Lalagkas and Melamed 2026). But, our previous design had a number of weaknesses: 1) It is limited in the data on drug, disease, and gene biology that can be integrated; 2) It relies on a linear design which is simple and interpretable but not flexible; 3) It relies on dimensionality reduction, creating unstable connections between drugs and disease genes. Here we present DraPhormer, a transformer based architecture that integrates molecular and genetics data along with language model representations of genes, drugs, and diseases. These representations allow us to greatly reduce the learning space, enabling robust learning of connections between drugs and disease genes explaining drug effects on disease. We expect this model will allow insight into the biology of drug effects on disease.

## Methods

### Transformer model details

DraPhormer aims to learn attention scores between drug and gene that explain the connection between a drug and phenotype. To this end, the model is based off of drug, gene, and phenotype representations. The embeddings for a drug are represented in a matrix *D* of dimension *d*, and for a gene we have embeddings in a matrix *G* of dimension *g*. Then, we must learn matrices *W*_*Q*_ ∈ *R*^*d*×*k*^ and *W*_*K*_ ∈ *R*^*g*×*k*^, each transforming to shared hidden dimension *k*.

Together they calculate the attention score: 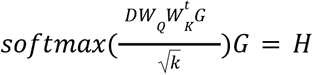. We call the output of this calculation, per drug, *drugRep*_*d*_ because it maps a drug to its representation in the space of disease genes. Finally, the distance between this *drugRep*_*d*_ and any phenotype, in terms of cosine similarity, is used to quantify the probability that the drug treats the phenotype.

### Preparation of molecular, genetic, and drug therapeutic effects data

We have previously described the preparation of the data for Draphnet. Briefly, we use genome-wide association study (GWAS) results to create a p-value for each gene and disease combination, summarizing the extent to which variants in impacting regulation of that gene are associated with risk of disease (Pividori et al. 2020). We denote this matrix as *P* with rows as each gene, and columns comprising hundreds of disease phenotypes. We match diseases to their UMLS codes (Bodenreider 2004) in order to obtain disease to drug information from SIDER (Kuhn et al. 2016). We obtain the ToxCast data from https://www.epa.gov/chemical-research/exploring-toxcast-data which comprises our matrix *D* linking each of 429 compounds to its effect on up to 1391 endpoints (though over 70% of the matrix is missing, corresponding to unmeasured drug-endpoint pairs).

Next, we obtain embedding representations for genes, drugs, phenotypes, and endpoints. We use DRKG to represent drugs (gnn4dr [2020] 2023). This is based off of a knowledge graph and trained to represent an embedding for each drug. For phenotypes, we use sapbert to represent each disease. Unlike DRKG, which sapbert is a language model. We feed the disease text name into the model and use the final token as the representation. We perform a similar process for the ToxCast endpoints. For genes we use BIOCONCEPTVEC-FASTTEXT, based on benchmark performance for retrieving disease genes (Zhong et al. 2025).

### Simulation setup and evaluation

Our simulation takes two steps. We simulate the matrix *P* randomly, connecting diseases to genes such that each disease connects to, on average, 5 genes. Then, we simulate the gene and disease phenotype embeddings such that genes linked similar sets of phenotypes share similar embeddings, and likewise for phenotypes. Then, we simulate the transformer matrices *W*_*Q*_ and *W*_*K*_, and we scale the learned drug to gene attention scores such that some genes-drug connections have probability greater than 0.1. Using the representation of each drug, *drugRep*_*d*_ we obtain the dot product between *drugRep*_*d*_ and the disease embedding vector. This value is again scaled linearly using simulated scale and additive factors, and then passed through a sigmoid to obtain a probability of a drug to disease connection. The probabilities are used for sampling binary simulated drug therapeutic uses using a bernoulli.

Then, given embedding vectors and the simulated drug to phenotype connections in the training set, we ask whether the model can recover the following from drugs in the test set: 1) the drug to phenotype connections, measured using the area under the precision recall curve; and 2) the drug to gene attention scores, measured using the spearman correlation between the simulated and recovered values.

## Results

### Design of a model to learn the drugs impacting disease genes

Our model assumes that a drug’s impact on a phenotype can be predicted as a combination of the drug’s impact on disease genes. To encode this logic into a machine learning model, we use the transformer architecture. The transformer model learns an attention score connecting each drug to a set of disease genes, then representing the drug as a linear combination of its connected disease genes. Finally, using the similarity between this drug representation and any phenotype, we predict whether the drug impacts the phenotype (Figure 1). This design allows all predictions of drug effect to be made in terms of the attention scores connecting drugs to disease genes. It also is flexible, allowing encoding information about drugs, genes, and phenotypes both using the embedding features of each, and by enriching the model with graph connections.

**Figure 1.**
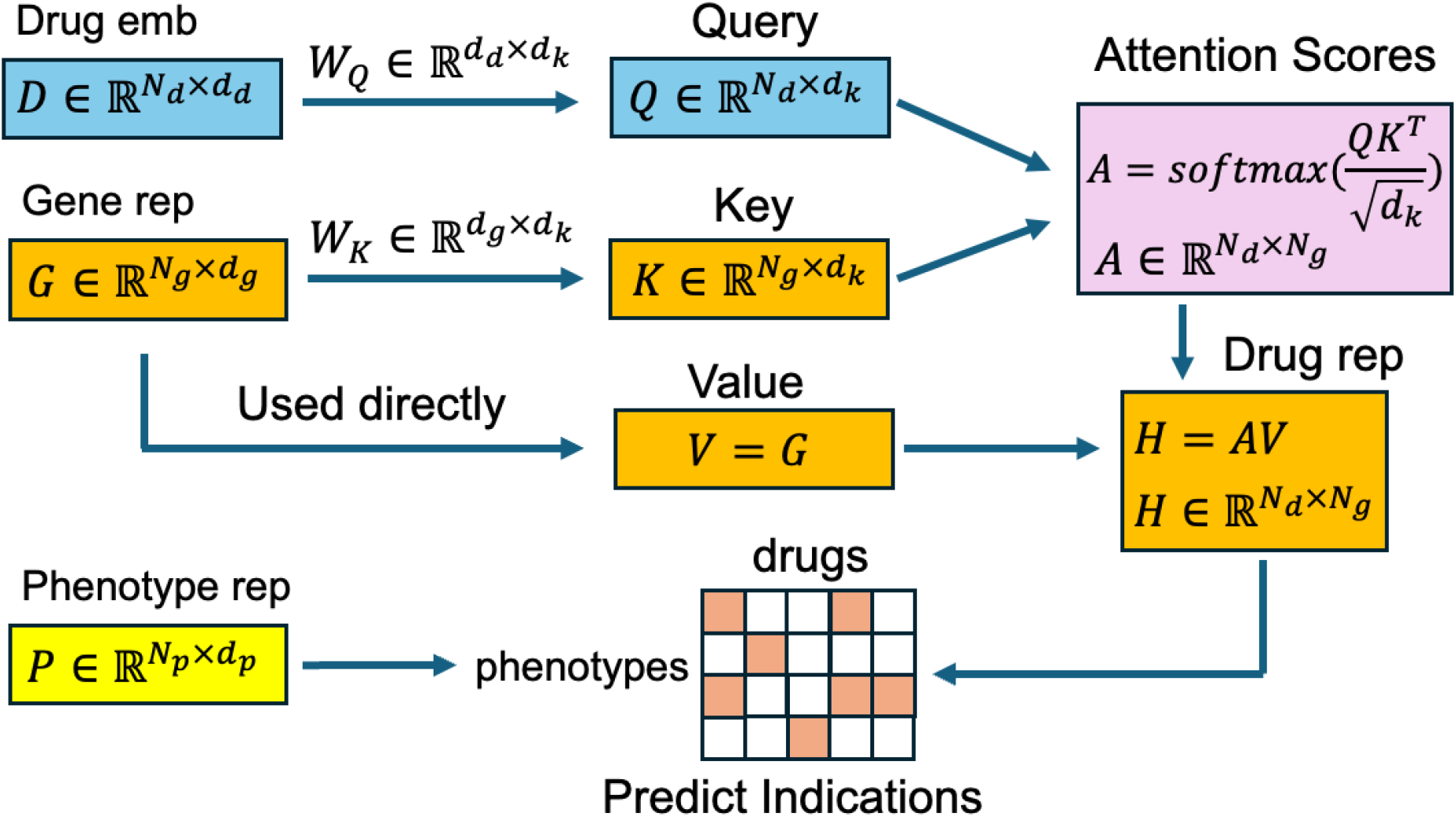
DraPhormer design.

### Simulation design and performance

We simulate data for DraPhormer to ask whether this architecture is able to learn the connections between drugs and disease genes that explain drug effects on disease phenotype. In our simulation, the generated data follows the hypothesized structure (see Methods). That is, drug-phenotype connections are simulated such that drugs treat a disease when they are connected to disease genes that are, in turn, connected to that disease phenotype.

Figure 2 shows that in our simulation, DraPhormer is able to recover not only the drug to disease predictions of therapeutic effects (Figure 2A), but also the drug to disease gene attention scores that explain these predictions (Figure 2B).

**Figure 2.**
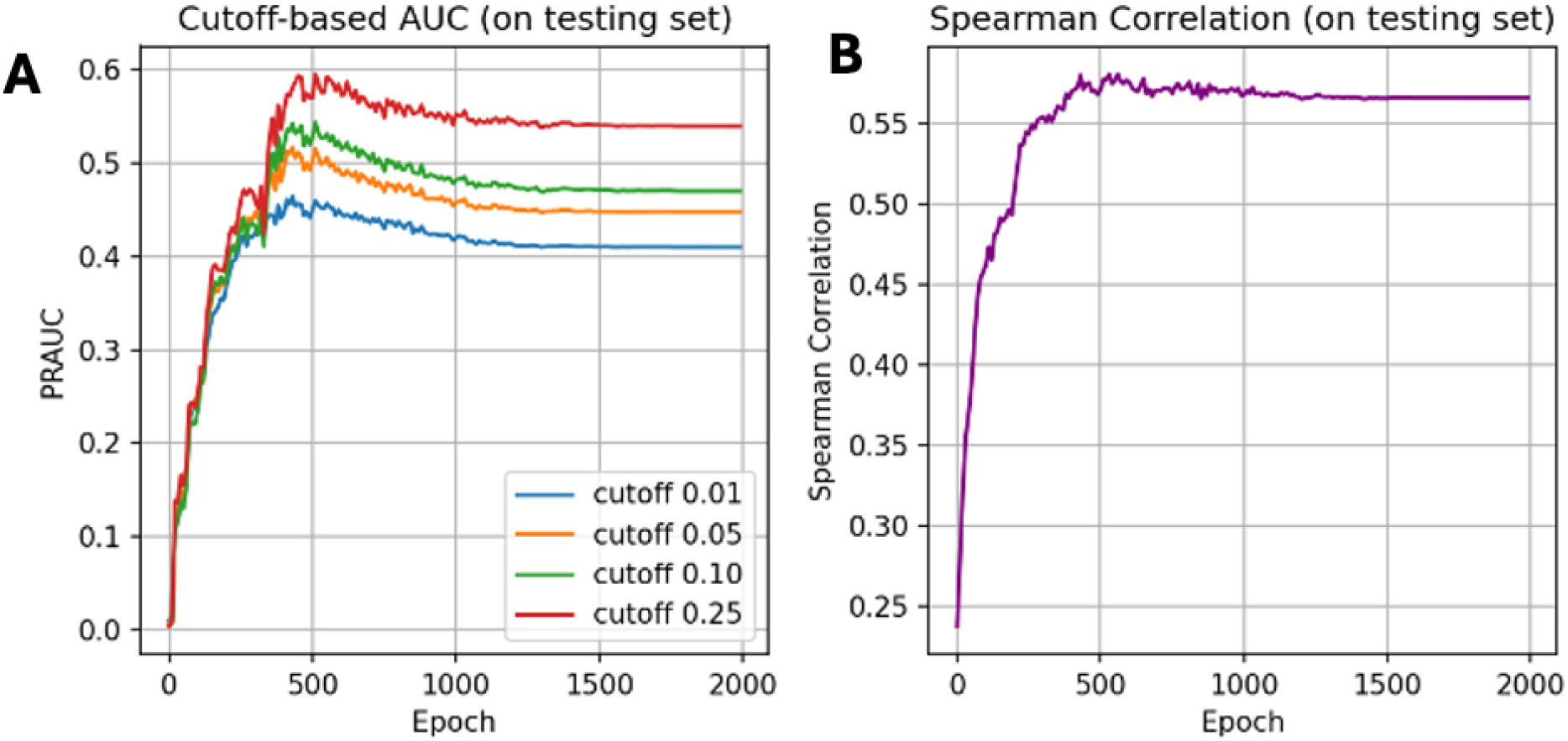
DraPhormer performance on the simulation test set in terms of recovering drug-disease connections (A), and in terms recovering drug-gene connections (B). **A.**Drug-disease connections are simulated as in Methods, where the simualated probability of a drug treating a disease is used to sample binary labels. The area under the precision recall curve tests various cutoffs of the probability. **B**. The simulated attention matrices yield an attention score for each drug-gene pair. We compare the simulated versus learned attention scores in terms of Spearman correlation.

**Figure 3.**
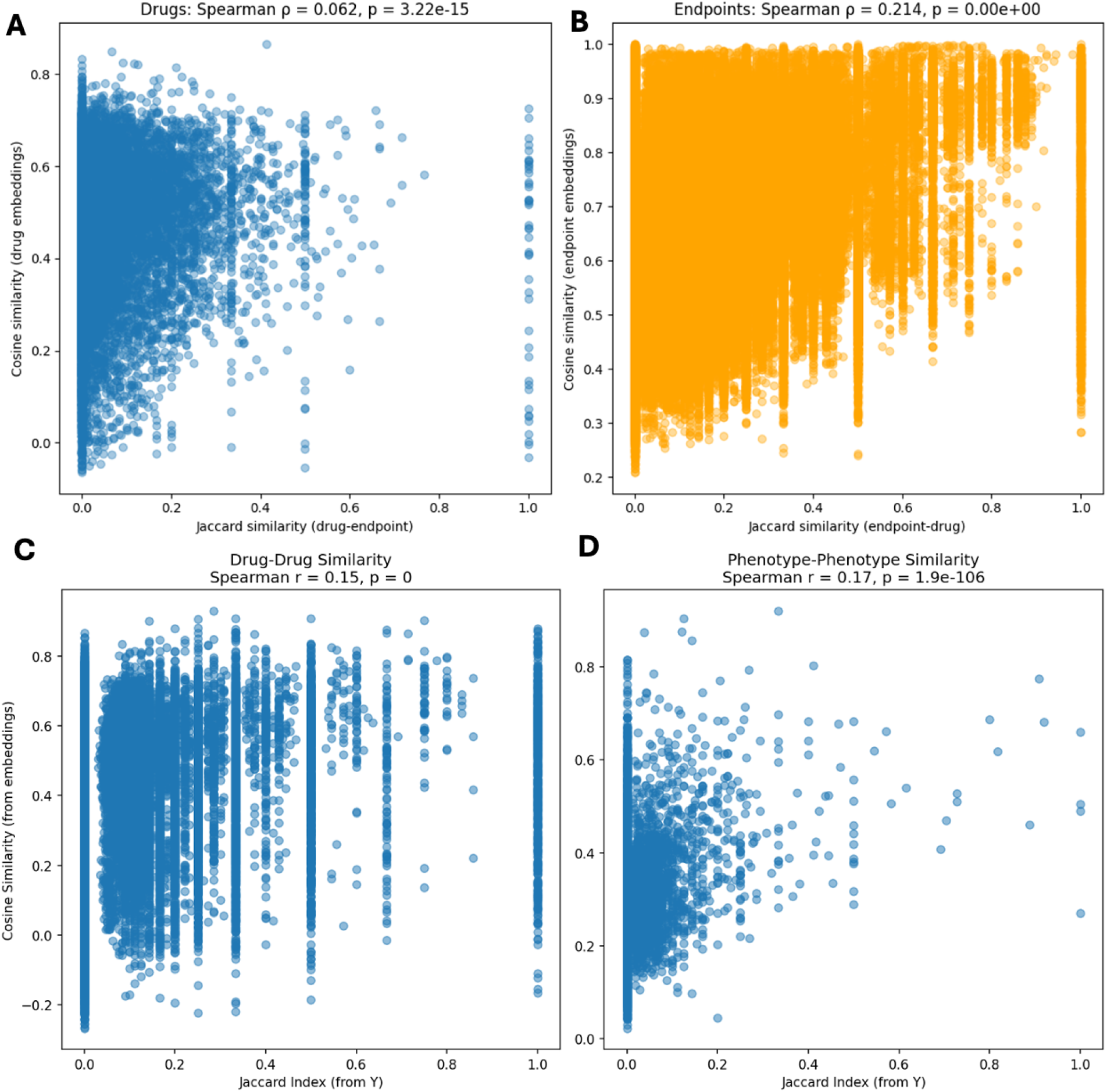
Evaluation of consistency of language model embeddings and molecular and clinical data. **A.**Drugs sharing more endpoint connections have more similar embeddings. **B**. Endpoints connected to more overlapping sets of drugs have more similar embeddings. **C**. Drugs sharing more therapeutic uses have more similar embeddings. **D**. Phenotypes sharing more treating drugs have more similar embeddings.

### Preparation of molecular data and language model representations for model training

Next, we must obtain representations for each gene, drug, and disease for model training. Importantly, although our simulation linked such representations to the disease genetics data, we do not know how the model can perform using real representations, as opposed to simulated features. We also obtain ToxCast representation of *in vitro* effects of drugs on disease-relevant molecular endpoints; results connecting drugs to disease genes from S-PrediXcan; and drug therapeutic or adverse effects on diseases from SIDER (see Methods).

We obtain embeddings from language models as described in Methods. Then, we ask whether drugs with similar embeddings share similar molecular profiles, as encoded in the drug to endpoint matrix (*D*, see Methods) and drug to phenotype matrix (*P*, see Methods). There is a correlation though much weaker than in our simulated data.

### Evaluation of model performance on recovering drug effects

We use embeddings derived from language models in place of our simulated embeddings, as described above. Then, we simulate the attention matrices and ask if we can recover the true (simulated) drug to gene attention scores. As shown in Figure 4, the model is able to recover the simulated signal, although the fidelity is much lower. For example, with full simulation, the connection between the drug and disease embeddings enabled learning of drug to gene connections with spearman correlation above 0.5 in the test set, while in this model, the correlation never exceets 0.25.

**Figure 4.**
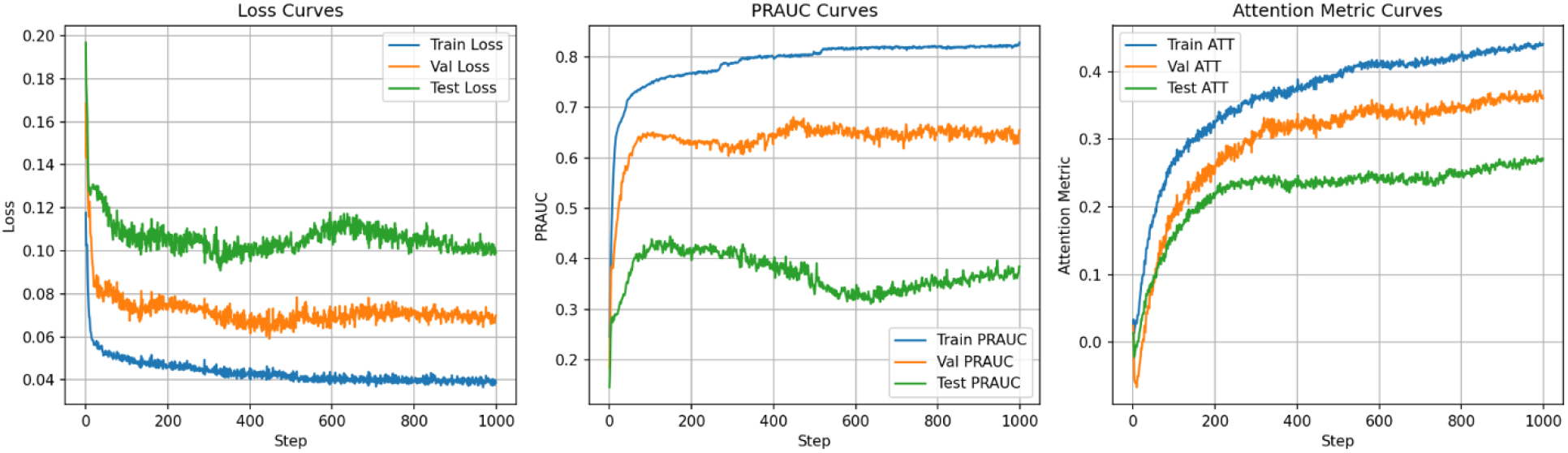
Model evaluation with simulated attention and real embeddings, in terms of loss, PRAUC, and correlation of attention scores.

## Discussion

While one approach to explainable machine learning uses post-hoc analysis to learn the importance of each feature to the prediction, here we embed explainability into the model design. We expect that each predicted effect of a drug on a disease occurs through drug to gene connections. A model that is not designed to predict drug to disease connections in this way is likely to use other features to make predictions. Therefore, a model where the architecture explicitly only allows predictions of drug to phenotype connections to occur via the impacts of drugs on disease genes can provide unique biological insight.

Our current model has many fewer parameters to learn than the previous Draphnet model, and also does not rely on dimensionality reduction, a major strength. As well, it can incorporate molecular data, including drug *in vitro* assays, together with disease genetics information, and embedding representations of genes, drug, and diseases encoded in language models. As the language models are trained incorporating biomedical literature and other human created sources, they have the potential to describe complex understanding of the biology of each entity.

Our model is not yet successful at learning to describe the biology of drug effects. This is due to the disparity between language models and disease genetic data. Our next steps are to fine tune the models to reconcile these representations. We will use external data on drug biology, similar to our previous work, to evaluate whether drugs with similar inferred disease genes share known biological effects. As well, we will compare all findings with the previous Draphnet model.

## Data Availability

All data is publicly available.

https://www.epa.gov/chemical-research/exploring-toxcast-data

http://sideeffects.embl.de/

https://zenodo.org/records/3685379

